# Diagnostic power of chest CT for COVID-19: to screen or not to screen

**DOI:** 10.1101/2020.05.18.20097444

**Authors:** K. De Smet, D. De Smet, I. Demedts, B. Bouckaert, T. Ryckaert, E. Laridon, B. Heremans, R. Vandenbulcke, S. Gryspeerdt, G.A. Martens

## Abstract

**Background:** chest CT is increasingly used for COVID-19 screening in healthcare systems with limited SARS-CoV-2 PCR capacity. Its diagnostic value was supported by studies with methodological concerns and its use is controversial. Here we investigated its potential to diagnose COVID-19 in symptomatic patients and to screen asymptomatic patients in a prospective study with minimal selection bias.

**Methods:** From March 19, 2020 to April 20, 2020 we performed parallel SARS-CoV-2 PCR and CT with categorization of COVID-19 suspicion by CO-RADS, in 859 patients with COVID-19 symptoms and 1138 controls admitted to the hospital for COVID-19 unrelated medical urgencies. CT-CORADS was categorized on a 5-point scale from 1 (very low suspicion) to 5 (very high suspicion). AUC under ROC curve were calculated in symptomatic versus asymptomatic patients to predict positive SARS-CoV-2 positive PCR and likelihood ratios for each CO-RADS score were used for rational selection of diagnostic thresholds.

**Findings:** CT-CORADS had significant (P<0.0001) diagnostic power in both symptomatic (AUC=0.891) and asymptomatic (AUC=0.700) patients hospitalized during SARS-CoV-2 peak prevalence. In symptomatic patients (41.7% PCR+), CO-RADS ≥ 3 detected positive PCR with high sensitivity (89.1%) and 72.5% specificity. In asymptomatic patients (5.3% PCR+), a CO-RADS score ≥ 3 detected SARS-CoV-2 infection with low sensitivity (45.0%) but high specificity (88.8%).

**Interpretation:** CT-CORADS has meaningful diagnostic power in symptomatic patients, supporting its application for time-sensitive triage. Sensitivity in asymptomatic patients is insufficient to justify its use as screening approach. Incidental detection of CO-RADS ≥ 3 in asymptomatic patients should trigger reflex testing for respiratory pathogens.

## Introduction

Chest CT can be used to determine the temporal disease stage of COVID-19 pneumonia and measure its severity ^1-3^. In the early stage of viral replication (day 0-4) ground-glass opacities are the predominant lesion. In the progressive stage (day 5-8), crazy paving patterns mark the increased recruitment of inflammatory cells to the lung interstitial space. Peak stage (day 10-13) is marked by consolidation due to tissue organization with fibrosis and diffuse alveolar damage. These radiological lesions are not specific for SARS-CoV-2 and are observed also in other viral pneumonia and non-infectious inflammatory lung diseases. In a pandemic setting, however, with high prevalence of SARS-CoV-2 infection and low prevalence of other viral lung infections, these non-specific changes harbor diagnostic power to diagnose COVID-19 in symptomatic patients or to screen presymptomatic infections as part of infection control measures. The reference method for COVID-19 diagnosis, SARS-CoV-2 PCR, is highly specific but its sensitivity is variable and might be as low as 70% in the early stage of infection ^4^ due to low viral loads and suboptimal sampling. In overwhelmed health care systems, limited PCR capacity and relatively slow turnaround times create bottle necks for efficient triage. Chest CT is increasingly used as fast and readily available alternative to diagnose or screen COVID-19 and with limited scientific scrutinization integrated in practice and guidelines ^5^. As outlined by Hope M. and Raptis C. et al ^6-8^, studies supporting the chest CT for COVID-19 screening showed major methodological concerns. First, most studies were underpowered with the exception of one study on 1014 symptomatic COVID-19 patients that reported a sensitivity of 97% at a low specificity of 25% ^3^. Second, all suffered from major selection biases by including only patients with COVID-19 symptoms and a 40%-50% a priori risk of SARS-CoV-2 infection. Third, all used binary scoring of CT without a standardized and reproducible definition of COVID-19-compatible CT. Taking into account the medical risk of unnecessary radiation exposure and a possible additional chain of viral transmission in insufficiently decontaminated scanners, the lack of high-quality data sparked a controversy ^8, 9^ leading to consensus statements by The Centers for Disease Control and Prevention, the American College of Radiology, and the Society of Thoracic Radiology and American Society of Emergency Radiology opposing the use of CT as diagnostic and screening tool for COVID-19 ^10,11^. In this report, we studied the diagnostic power of chest CT as compared to SARS-CoV-2 PCR in 859 patients admitted with WHO-listed COVID-19 symptoms and its potential as a COVID-19 screening method in 1138 subjects without COVID-19 symptoms admitted for other urgent medical needs. Chest CT images were scored using the Dutch CO-RADS (COVID-19 Reporting and Data System) classification system, allowing categorical assessment of the level of suspicion ^12^. We attributed likelihood ratios to each CO-RADS score in symptomatic and asymptomatic subjects, allowing the rational selection of diagnostic thresholds and better understanding of its real diagnostic value as function of the overall SARS-CoV-2 infection prevalence.

## Methods

### Patients

This is a prospective cohort study on 1997 consecutive patients admitted to AZ Delta General Hospital in Roeselare, Belgium from March 19, 2020 to April 20, 2020: 859 subjects (male/female: 443/416 Age, y, median (IQR): 70 (52-81)) with symptoms matching the COVID-19 case definition as specified by the World Health Organization (WHO) interim guidance of February 27, 2020 ^13^ (further referred to as ‘symptomatic patients’) and 1138 subjects (male/female: 588/550, Age, y, median (IQR): 68 (52-80)) without clinical suspicion of COVID-19 (further referred to as ‘asymptomatic patients’) admitted for COVID-19-unrelated urgent medical needs (surgical and endoscopic procedures, psychiatric illness and gerontology) who were screened by chest CT and SARS-CoV-2 PCR as part of the medical board-approved triage policy for SARS-CoV-2 quarantining. The study was approved by the AZ Delta Institutional Review Board with a waiver of informed consent from study participants considering the study is based on secondary analysis of existing data (Clinical Trial Number: B1172020000008)

### Procedures

On admission all subjects received a chest CT (detailed scanning protocol in Online supplement) with structured reporting by consensus evaluation of various indicators: temporal disease stage 1 to 3 of viral pneumonia, the extent of lung involvement by estimated residual aerated lung tissue on a scale of 1 to 5 in the 5 lobes and CO-RADS (COVID-19 Reporting and Data Systems). In this study, we restrict analysis to CO-RADS, since this categorical assessment scheme was specifically designed for diagnostic use. It grades the suspicion of COVID-19 pneumonia on a scale of 1 (very low level of suspicion) to 5 (very high level of suspicion) with CO-RADS 3 representing equivocal CT findings^12^.SARS-CoV-2 PCR was done with multiplex Real-time PCR for E/N/RdRP genes using Allplex™ 2019-nCoV assay (Seegene Inc, Seoul, Korea) on nasopharyngeal swabs.

### Statistical analysis

The diagnostic power of categorical CT-assessment by CO-RADS classification (CT-CORADS) was evaluated by calculating area (AUC) under the receiver operating characteristics (ROC) curve compared to SARS-CoV-2 PCR positivity. Likelihood ratios (LR, 95%CI) were calculated for each CO-RADS score in the symptomatic versus the asymptomatic group and visualized in diagrams of pre/post-test probability according to Bayes’ theorem. Statistical analyses were performed using MedCalc (version 12.2.1, Belgium) and considered significant if P value was less than .05.

## Results

### Diagnostic power in symptomatic patients

A total of 859 patients were admitted with WHO-listed symptoms of COVID-19 pneumonia: 443 males (median age 71 years, IQR 54-80 years) and 416 females (median age 68 years, IQR 51-82 years). On admission patients received combined chest CT for CORADS scoring (CT-CORADS) and SARS-CoV-2 PCR (Table 1). Overall prevalence of SARS-CoV-2 infection in symptomatic patients was 41.7%. In symptomatic patients with CO-RADS 5, 89.4% were PCR+ as compared to only 8.6% PCR+ cases in symptomatic patients with CO-RADS 1. ROC analysis confirmed the significant diagnostic power (P<0.001) of CT-CORADS with AUC = 0.891 (95%CI 0.868-0.911) to predict SARS-CoV-2 PCR-positivity (Fig. 1A). Next we calculated likelihood ratios (LR) for each CO-RADS score in symptomatic patients (Table 1): CORADS 1, 2 and also the ‘equivocal’ score CORADS 3 (LR=0.34, 95%CI 0.20-0.59) all decreased the odds of PCR positivity. CO-RADS 4 did not further increase post-test probability. CO-RADS 5 strongly, however, increased the odds of a positive PCR (LR=11.83 95%CI 8.47-16.53) (Fig. 1B). A CO-RADS 5 score in symptomatic patients identified SARS-CoV-2 PCR positivity with a sensitivity of 77.9% (95%CI 73.3-82.1) at high specificity of 93.4% (95%CI 90.9-95.4) and high overall accuracy of 87.0% (95%CI 84.5-89.1). Dichotomization of suspected CT at CO-RADS ≥ 4 and ≥ 3 increased sensitivity to 84.3% (95%CI 80.8-88.5) and 89.1% (95%CI 85.4-92.1) at a specificity of 84.8% (95%CI 68.3-76.3) and 72.5% (95%CI 68.376.3) respectively (Table 1). The diagram in Fig.1B plots the associated shift from pre-test probability (overall prevalence of positive PCR) to post-test probability of SARS-CoV-2 infection in individual patients as function of CO-RADS score.

**Table 1.**
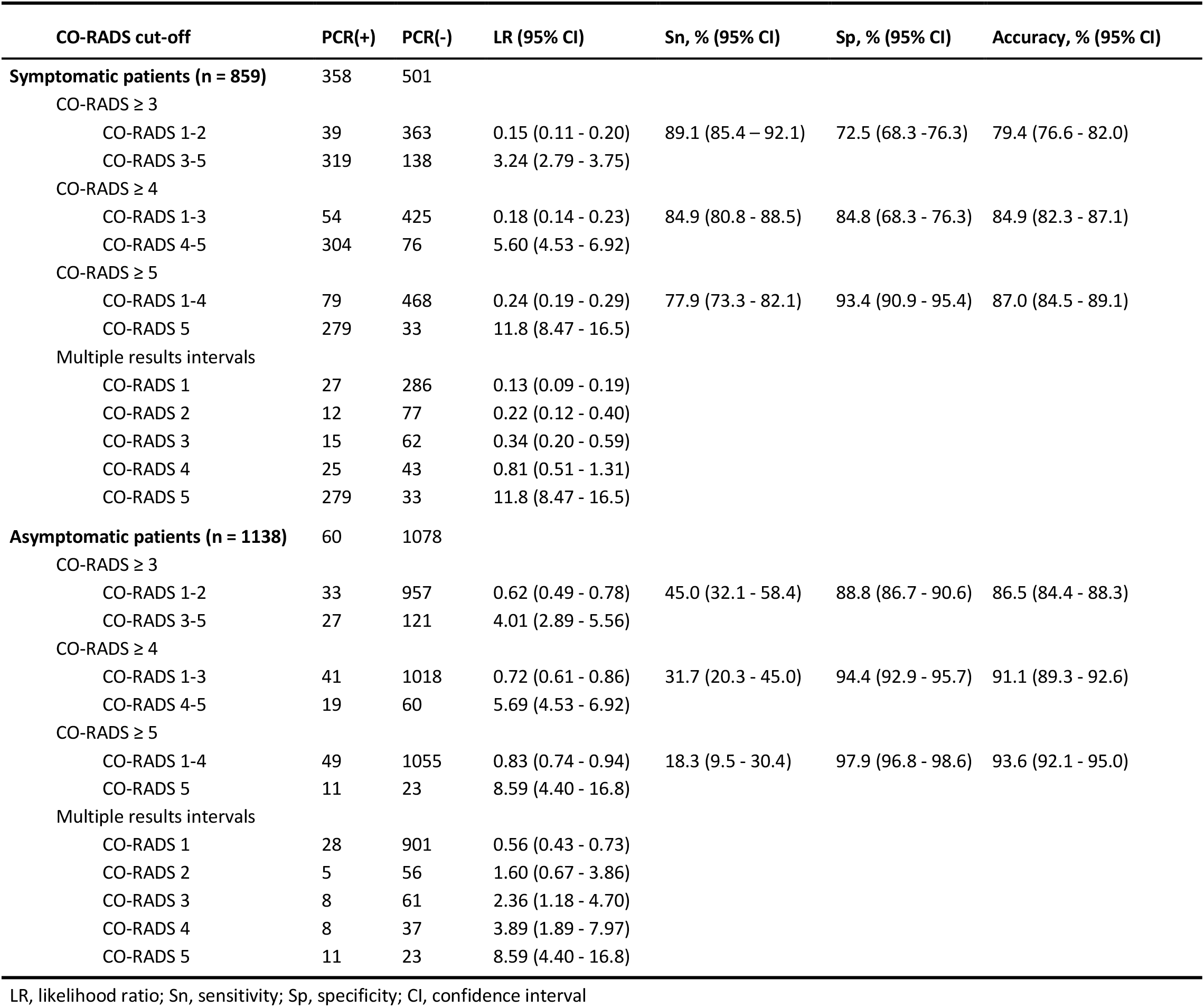
Diagnostic performance of CO-RADS for symptomatic and asymptomatic setting at different CO-RADS cut-offs and multiple result intervals. The table shows the distribution of asymptomatic and symptomatic patients over multiple result intervals (CO-RADS score 1 to 5) and various possible dichotomization approaches, with their associated number of positive/negative PCR tests and the associated likelihood ratios (LR, 95% confidence interval) to predict positive PCR. The right columns indicate the associated sensitivity (Sn), specificity (Sp) and accuracy and 95% confidence intervals.

| CO-RADS cut-off | PCR(+) | PCR(-) | LR (95% CI) | Sn, % (95% CI) | Sp, % (95% CI) | Accuracy, % (95% CI) |
| --- | --- | --- | --- | --- | --- | --- |
| **Symptomatic patients (n = 859)** | 358 | 501 |  |  |  |  |
| CO-RADS $\geq 3$ | | | | | | |
| CO-RADS 1-2 | 39 | 363 | 0.15 (0.11 - 0.20) | 89.1 (85.4 - 92.1) | 72.5 (68.3 - 76.3) | 79.4 (76.6 - 82.0) |
| CO-RADS 3-5 | 319 | 138 | 3.24 (2.79 - 3.75) |  |  |  |
| CO-RADS $\geq 4$ | | | | | | |
| CO-RADS 1-3 | 54 | 425 | 0.18 (0.14 - 0.23) | 84.9 (80.8 - 88.5) | 84.8 (68.3 - 76.3) | 84.9 (82.3 - 87.1) |
| CO-RADS 4-5 | 304 | 76 | 5.60 (4.53 - 6.92) |  |  |  |
| CO-RADS $\geq 5$ | | | | | | |
| CO-RADS 1-4 | 79 | 468 | 0.24 (0.19 - 0.29) | 77.9 (73.3 - 82.1) | 93.4 (90.9 - 95.4) | 87.0 (84.5 - 89.1) |
| CO-RADS 5 | 279 | 33 | 11.8 (8.47 - 16.5) |  |  |  |
| Multiple results intervals |  |  |  |  |  |  |
| CO-RADS 1 | 27 | 286 | 0.13 (0.09 - 0.19) |  |  |  |
| CO-RADS 2 | 12 | 77 | 0.22 (0.12 - 0.40) |  |  |  |
| CO-RADS 3 | 15 | 62 | 0.34 (0.20 - 0.59) |  |  |  |
| CO-RADS 4 | 25 | 43 | 0.81 (0.51 - 1.31) |  |  |  |
| CO-RADS 5 | 279 | 33 | 11.8 (8.47 - 16.5) |  |  |  |
| **Asymptomatic patients (n = 1138)** | 60 | 1078 |  |  |  |  |
| CO-RADS $\geq 3$ | | | | | | |
| CO-RADS 1-2 | 33 | 957 | 0.62 (0.49 - 0.78) | 45.0 (32.1 - 58.4) | 88.8 (86.7 - 90.6) | 86.5 (84.4 - 88.3) |
| CO-RADS 3-5 | 27 | 121 | 4.01 (2.89 - 5.56) |  |  |  |
| CO-RADS $\geq 4$ | | | | | | |
| CO-RADS 1-3 | 41 | 1018 | 0.72 (0.61 - 0.86) | 31.7 (20.3 - 45.0) | 94.4 (92.9 - 95.7) | 91.1 (89.3 - 92.6) |
| CO-RADS 4-5 | 19 | 60 | 5.69 (4.53 - 6.92) |  |  |  |
| CO-RADS $\geq 5$ | | | | | | |
| CO-RADS 1-4 | 49 | 1055 | 0.83 (0.74 - 0.94) | 18.3 (9.5 - 30.4) | 97.9 (96.8 - 98.6) | 93.6 (92.1 - 95.0) |
| CO-RADS 5 | 11 | 23 | 8.59 (4.40 - 16.8) |  |  |  |
| Multiple results intervals |  |  |  |  |  |  |
| CO-RADS 1 | 28 | 901 | 0.56 (0.43 - 0.73) |  |  |  |
| CO-RADS 2 | 5 | 56 | 1.60 (0.67 - 3.86) |  |  |  |
| CO-RADS 3 | 8 | 61 | 2.36 (1.18 - 4.70) |  |  |  |
| CO-RADS 4 | 8 | 37 | 3.89 (1.89 - 7.97) |  |  |  |
| CO-RADS 5 | 11 | 23 | 8.59 (4.40 - 16.8) |  |  |  |

LR, likelihood ratio; Sn, sensitivity; Sp, specificity; CI, confidence interval

**Fig. 1:**
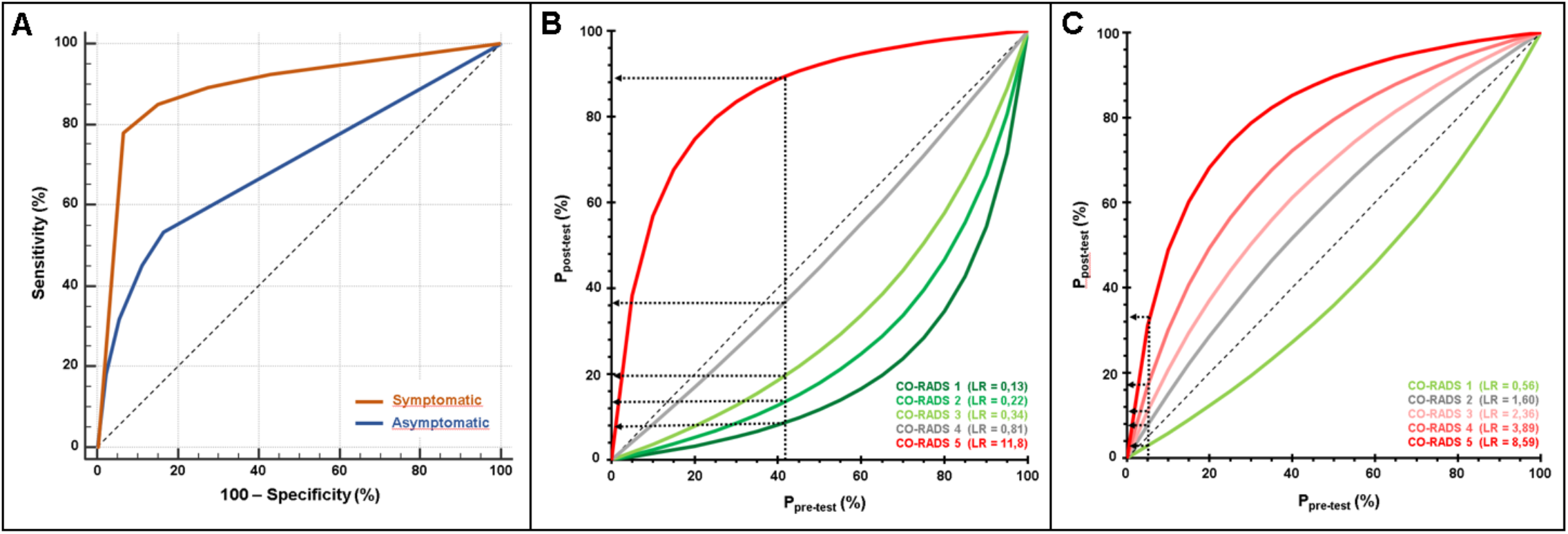
Diagnostic power of CT-CORADS scoring in patients with and without COVID-19 symptoms. **A**. The area under the receiver operating characteristics curve (AUC) of CT-CORADS to predict a positive SARS-CoV-2 PCR result in symptomatic (red line) and asymptomatic (blue line). The diagonal dashed line indicates no discrimination. **B**. Post-test probability of positive PCR as function of the pre-test probability for different likelihood ratios (LR) associated with the indicated CO-RADS score in 859 symptomatic patients. The arrow indicates the pre-test probability as determined by overall prevalence of positive PCR (41.7%) in this cohort. C. Post-test probability of positive PCR as function of the pre-test probability for different likelihood ratios (LR) associated with the indicated CO-RADS score in 1138 asymptomatic patients. The arrow indicates the pre-test probability as determined by overall prevalence of positive PCR (5.2%) in this cohort.

### Screening potential of chest-CT in asymptomatic patients in a SARS-CoV-2 pandemic setting

From March 19, 2020 to April 20, 2020 a total of 1138 subjects were admitted for urgent medical needs (surgery, endoscopic procedures, psychiatric illness and gerontology) unrelated to WHO-listed COVID-19 symptoms (‘asymptomatic patients’): 588 males (median age 66 years, IQR 53-78 years) and 550 females (median age 70 years, IQR 50-82 years). As part of institutional infection control guidance, all admitted patients received a combi-screening with CT-CORADS for first rapid triage and a SARS-CoV-2 PCR with turnaround time below 24 hours. Prevalence of SARS-CoV-2 PCR-positivity in asymptomatic patients was 5.3% (60/1138). 6.9% of asymptomatic patients showed a CO-RADS score of 4 (high suspicion) or 5 (very high suspicion), 87.0% showed a CO-RADS score ≤ 2 with low to very low suspicion of COVID-19 (Table 1). ROC analysis indicated that CT-CORADS in asymptomatic patients had diagnostic power (P<0.001) to predict SARS-CoV-2 PCR-positivity with AUC = 0.700 (95%CI 0.672-0.726) (Fig. 1A), albeit significantly less than in symptomatic patients. The percentage of PCR-positive cases was 3.0%, 8.2%, 11.6%, 17.8% and 32.4% in CO-RADS 1, 2, 3, 4 and 5, respectively. Analysis of likelihood ratios (Table 1, Fig. 1C) indicated that only CO-RADS 1 could slightly lower the odds of a positive PCR (LR=0.56, 95%CI 0.43-0.73), that CO-RADS 2 had no diagnostic meaning with the 95% CI encompassing LR=1 and that CO-RADS 3 and higher, chest CT increased the odds of a positive PCR, resulting in a positive shift from pre- to post-test probability (Fig. 1C). In particular CO-RADS 5 had strong diagnostic power in asymptomatic patients, with LR =8.59 (95%CI 4.40-16.79), predicting SARS-CoV-2 infection at high specificity of 97.9% (95%CI 96.8-98.6) but low sensitivity of 18.3% (95%CI 9.5-30.4). Dichotomization of suspected CT at CO-RADS ≥ 4 preserved a high specificity of 94.4% (95%CI 92.9-95.7) but its sensitivity of 31.7% (95%CI 20.3-45.0) was still too low to advocate its use as a screening tool.

## Discussion

CO-RADS was developed by the COVID-19 Standardized Reporting Working Group of the Dutch Radiological Society as a categorical system to score the level of suspicion for COVID-19 pulmonary involvement. In a proof-of-concept study on 105 patients with COVID-19 symptoms and a prevalence of SARS-CoV-2 PCR-confirmed infection of 50.5%, CT-CORADS show strong diagnostic power with AUC under the ROC curve of 0.91 (95% CI 0.85-0.97) ^12^. Our study confirms this diagnostic power with similar AUC of 0.891 (95%CI 0.868-0.911) on a larger cohort of 859 symptomatic patients with a prevalence of 41.7% SARS-CoV-2 infections, indicating robustness of the scoring system.

A novelty in our approach was the attribution of likelihood ratios to each CO-RADS score, allowing rational selection of possible dichotomizations as function of the clinically desired sensitivity/specificity. In a high prevalence setting - symptomatic patients in a pandemic setting - a CORADS ≥ 3 achieves acceptable sensitivity of 89.1% and specificity of 72.5% to serve as pre-triage. CORADS 5 was particularly powerful and strongly increased the odds of a positive PCR from 41.7% to 89.4% and could thus serve as alternative triage in emergency settings with bottlenecks in PCR testing.

This study is the first to investigate the diagnostic power of chest CT in a large cohort of 1138 diseased control patients without COVID-19 symptoms. In these ‘asymptomatic patients’, prevalence of SARS-CoV-2 PCR-confirmed infection was only 5.3%, in line with the reported secondary attack rate at population level of 6.6% during the exponential phase ^14^. This allowed us to investigate the controversial issue of upfront screening for COVID-19 by chest CT in subjects without COVID-19 symptoms. The answer is negative: though CT-CORADS has diagnostic power even in asymptomatic patients, and though the odds of a positive PCR increased with increasing CO-RADS score above 1, various dichotomization scenarios failed to reach the required high sensitivity (around 90%) for a screening test, e.g. only 31.7% sensitivity for CO-RADS score ≥ 4. While its sensitivity is too low to classify as a screening test, considering the additional medical risk of CT, the corresponding specificity was excellent, e.g. 94.4% in asymptomatic patients with CO-RADS ≥ 4. Any such result, if incidentally obtained in an asymptomatic patient undergoing CT for any other reason, should prompt targeted reflex testing by SARS-CoV-2 PCR or - if other respiratory viruses are simultaneously prevalent - syndromic panel-based PCR testing for respiratory pathogens and exclusion of non-infectious inflammatory lung diseases.

As compared by prior studies advocating the standard and broad use of chest CT for screening and triage of COVID-19 ^3,15^ our study has three methodological strengths. First, it is sufficiently powered. Second, by its prospective design with up-front distinction between, patients with or without COVID-19 symptoms, it does not suffer from selection bias by only studying patients with very high a priori risk of disease. Third, it uses a robust and well specified categorical CT interpretation, rather than an undefined dichotomous scoring.

The main limitation of our study is that it was conducted in the pandemic phase of SARS-CoV-2 infection, in a time frame with low prevalence of other respiratory viral infections such as influenza that can induce similar radiological abnormalities. In the influenza season, the diagnostic power of CORADS 5 for SARS-CoV-2 infection will decrease. In clinical practice, however, this does not argue against its medical utility, since a CO-RADS score of 5 is always an informative finding that should trigger targeted reflex testing for respiratory pathogens and differentiation between infectious versus non-infectious inflammatory conditions. In addition to its diagnostic value, chest CT is evidently also useful to assess the overall severity of pulmonary involvement (number of affected lobes and residual amount of well-aerated functional tissue) in COVID-19, and provides a direct view on the temporal evolution of SARS-CoV-2 infection as proxy for its immunological stage. This might harbor prognostic value, outside the scope of present analysis. Finally, chest CT allows the detection of other medical conditions with similar symptoms as COVID-19 such as bacterial pneumoniae, pleural effusion, lung cancer, pneumothorax and cardiac failure.

In conclusion, our data provide clear guidance for rational use of chest CT in COVID-19 triage during a pandemic phase. CT with structured CO-RADS scoring has strong diagnostic power for COVID-19 pneumonia. In symptomatic patients, a CO-RADS ≥ 3 serves as sensitive and rapid triage tool, in addition to being useful to determine disease stage and severity. In patients without COVID-19 symptoms, chest CT screening is not recommended due to low sensitivity but any incidental finding of CO-RADS scores ≥ 3 has high specificity for viral infection and should trigger reflex testing for respiratory pathogens.

## Data Availability

Source data discussed in the paper are available on email request to the guarantor of this study.

## Author contribution statement and data sharing statement

Study design: KDS, DDS, GM, SG. Statistical data analysis, lead: DDS. Statistical analysis, supportive: KDS, GM. Data interpretation: KDS, DDS, GM, SG, ID. Data collection: KDS, SG. Manuscript preparation, lead: GM, KDS, DDS. Manuscript, supportive: SG, BB, TR, EL, BH, RV, ID. GM is guarantor of the study.

## Acknowledgements

The authors thank H. Verelst, F. Trenson, L. Cruyt and J. De Melio for expert interpretation of chest CT and data contribution to consensus CO-RADS scoring.

